# Abnormal Blood Pressure Among Individuals Evaluated for Latent Tuberculosis Infection in a U.S. Public Health Tuberculosis Clinic

**DOI:** 10.1101/2024.06.24.24309439

**Authors:** Trevor M. Stantliff, Argita D. Salindri, Rocio Egoavil-Espejo, Ashton D. Hall, Laura Medina-Rodriguez, Kavya Patel, Matthew J. Magee, Elaine M. Urbina, Moises A. Huaman

## Abstract

Tuberculosis infection (LTBI) has been associated with increased cardiovascular risks. We aimed to characterize abnormal blood pressure (BP) readings in individuals with LTBI. We conducted a retrospective study of adults with LTBI presenting for their initial medical visit at a large midwestern US public health clinic between 2019-2020. Abnormal BP was defined as having a systolic BP≥ 130mmHg and/or a diastolic BP≥ 80mmHg. Of 310 individuals with LTBI, median age was 36 years (interquartile range 27 – 48), 34% were male, 64% non-US-born; 58 (18.7%) were previously diagnosed with hypertension. The prevalence of any hypertension (i.e., had a history of hypertension and/or an abnormal BP reading) was 64.2% (95% confidence interval 58.7 – 69.4). Any hypertension was independently associated with older age, male sex, higher body mass index, and Black race. In conclusion, any hypertension was present in over half of adults evaluated for LTBI in our clinic. Established hypertension risk factors were also common among this group, suggesting that individuals with LTBI could benefit from clinical and public health interventions aiming to reduce the risk of future cardiovascular events.

---

Despite the steady decline in active TB cases in the United States, the prevalence of latent tuberculosis infection (LTBI) has remained largely unchanged at 5% for the past three decades and disproportionately affects immigrant populations. (1) While the risk of progression of LTBI into active TB disease remains a major concern for TB surveillance and control programs, there is a growing appreciation of the interplay between LTBI and non-communicable diseases. For instance, recent studies reported that individuals with LTBI have increased rates of obstructive coronary artery disease, myocardial infarction, and diabetes mellitus incidence compared to their non-LTBI counterparts. (2-4)

LTBI has been linked with systemic inflammation and immune activation, which may contribute to increased cardiovascular risk including developing hypertension. (5, 6) Monocytes from individuals with LTBI exhibit pro-inflammatory alterations, including enhanced production of interleukin (IL)-6 and tumor necrosis factor (TNF)-α and increased expression of CX3CR1, a chemokine receptor implicated in monocyte migration, endothelial dysfunction, and atherosclerosis formation. (5, 7)

A recent analysis of the National Health and Nutrition Examination Survey cohort (NHANES) reported a high prevalence of hypertension (58.5%), a key risk factor for cardiovascular diseases, among civilian, non-institutionalized individuals with LTBI in the United States. (8) However, little is known whether hypertension/abnormal blood pressure (BP) (i.e., any hypertension) is more common among those with LTBI presenting to public health clinics that manage LTBI. In the present study, we aimed to estimate the prevalence of any hypertension and to determine if traditional risk factors for hypertension are similar among persons presenting for LTBI care in a real-world public health TB clinic setting. We also explored the relationship between quantitative measures of interferon-γ release assays (IGRA) and systolic and diastolic blood pressure.

We conducted a retrospective cohort study of adults (≥18 years old) who were referred to the Hamilton County Public Health (HCPH) clinic, a large midwestern US public health clinic located in Cincinnati, Ohio, for LTBI evaluation between 2019 and 2020. Eligible study participants included those referred to HCPH from community, private, and academic outpatient clinics and were determined to have LTBI. Individuals were defined as having LTBI based on a positive IGRA or tuberculin skin test (TST), negative chest-X-ray findings for active TB, and a negative clinical evaluation by a medical provider at HCPH. We excluded individuals with history of active tuberculosis or previous LTBI treatment. The initial study utilizing this cohort was designed to evaluate the effects of COVID-19 pandemic restrictions in the LTBI cascade of care, and those results have been presented elsewhere. (8) For the present analyses, we included sociodemographic and clinical data from all individuals who had available information on hypertension status and BP readings at their initial LTBI visit at HCPH.

We defined history of hypertension by patient’s self-report of prior diagnosis by healthcare providers and documented during the HCPH clinical encounter. BP at HCPH was measured while the patient was sitting and still, according to primary care guidelines. BP was recorded as systolic and diastolic measurements in mmHg. Abnormal BP was defined by having a systolic BP ≥ 130 mmHg, or a diastolic BP ≥ 80 mmHg. Our primary study outcomes, any hypertension, was defined as having either a history of hypertension or abnormal BP levels at initial visit.

Demographic, social risk factors, and clinical characteristics including tobacco use, intravenous (IV) drug use, history of homelessness and incarceration, current medications, as well as BCG vaccination status were determined by chart reviews. Body mass index (BMI) was calculated and classified with BMI <18.5 kg/m^2^ considered “underweight”, BMI 18.5 – 24.9 kg/m^2^ considered “normal” and BMI ≥25 kg/m^2^ considered “overweight/obese.”

We entered and managed data collected for this project in an online REDCap database hosted at the University of Cincinnati. We used Wilcoxon rank-sum tests to compare continuous variables between hypertension and non-hypertension groups. Similarly, we used Chi-square or Fisher’s exact tests to compare the distribution of categorial predictors. We performed robust Poisson regression models to estimate the prevalence ratios (PRs) comparing across different individuals’ characteristics and the 95% confidence interval (CI). Backward elimination technique was used to determine factors predictive of any hypertension. We also performed a subset analysis among individuals with quantitative IGRA measures to determine whether IGRA quantitative measures are associated with systolic and diastolic BP levels. We performed linear regression models to explore the relationship between 1) nil count, 2) TB antigen 1-nil, 3) TB antigen 2-nil, and 4) mitogen-nil values and systolic and diastolic BP. We reported beta estimates and corresponding 95% confidence intervals (95%CI) as well as R^2^ values to describe how well quantitative IGRA measures describe the observed systolic and diastolic BP data. All analyses were performed using Stata software (v12.0; StataCorp, College Station, TX) and R Statistical Software (v4.3.1; R Core Team 2023) with two-sided p-value <0.05 considered significant in all analyses.

Of the 312 individuals with LTBI presenting to the public health TB clinic for an initial visit with the medical provider, 310 (99.4%) had a hypertension status and recorded systolic and diastolic BP in their chart. Of 310 individuals with available BP readings, median age was 36 years (IQR, 27 – 48), 34% were male, 64% were non-US-born. Of these, 192 (61.9%) patients had an elevated BP (systolic BP ≥ 130mmHg or diastolic BP ≥ 80mmHg) (Supplementary Figure S1). Of the 192 patients with an elevated BP reading, only 51 (26.6%) had an established diagnosis of hypertension per history, while 141 others (73.4%) were newly diagnosed with elevated systolic or diastolic BP. There were seven patients with known history of hypertension who did not present with elevated BP at their initial visit. Thus, a total of 199 of 310 patients (64.2%, 95%CI 58.7 – 69.4) had either a known diagnosis of hypertension and/or an abnormal BP reading during their initial visit and were defined as the group of interest for downstream analyses.

Table 1 shows the characteristics of the study population stratified by BP status. The median of age among those with LTBI and any hypertension (median=41, interquartile range [IQR] 30 – 52) were significantly higher compared to those without hypertension (median=29, IQR 25 – 39, median difference=12 years, p-value <0.001). In the unadjusted model, the prevalence of hypertension was significantly higher among older age group (crude prevalence ratio [cPR]=1.46, 95%CI 1.22 – 1.74 for those aged 40 – 59 years and cPR=1.78, 95%CI 1.50 – 2.12 for those aged ≥60 years), male (cPR 1.31, 95%CI 1.12 – 1.54), Black race (cPR=1.53, 95%CI 1.28 – 1.83), with BMI ≥25kg/m^2^ (cPR=1.47, 95%CI 1.17 – 1.85), current/former smokers (cPR=1.36, 95%CI 1.16 – 1.59), and those with diabetes mellitus (cPR=1.38, 95%CI 1.14 – 1.67) (Table 1). The prevalence of hypertension among foreign born was 0.65 times the prevalence among US-born (95%CI 0.56 – 0.76). In the final multivariable model, the adjusted prevalence of hypertension remained significantly higher among those aged ≥60 years (adjusted prevalence ratio [aPR]=1.63, 95%CI 1.03 – 2.48), Black race (aPR=1.48, 95%CI 1.11 – 1.97), and those with BMI ≥25kg/m^2^ (aPR=1.47, 95%CI 1.04 – 2.10).

**Table 1:** Characteristics of individuals evaluated for latent tuberculosis at a large midwestern US public health clinic according to blood pressure status at initial visit (n=310).

| Participants<br>Characteristics | Total<br>population<br>(n=310) | No HTN/Abnormal<br>BP at initial visit<br>n (%) = 111 (35.8) | HTN/Abnormal BP<br>at initial visit<br>n (%) = 199 (64.2) | Measure of Association |  |
| --- | --- | --- | --- | --- | --- |
|  |  |  |  | Crude PR (95% CI) | Adjusted PR* (95% CI) |
| Age Group |  |  |  |  |  |
| 16 – 39 | 176 (56.8) | 84 (47.7) | 92 (52.3) | Reference | Reference |
| 40 – 59 | 105 (33.9) | 25 (23.8) | 80 (76.2) | **1.46 (1.22 – 1.74)** | 1.31 (0.97 – 1.78) |
| 60+ | 29 (9.4) | 2 (6.9) | 27 (93.1) | **1.78 (1.50 – 2.12)** | **1.63 (1.03 – 2.48)** |
| Sex |  |  |  |  |  |
| Female | 205 (66.1) | 86 (42.0) | 119 (58.0) | Reference | Reference |
| Male | 105 (33.9) | 25 (23.8) | 80 (76.2) | **1.31 (1.12 – 1.54)** | 1.28 (0.95 – 1.71) |
| Country of Birth |  |  |  |  |  |
| US-born | 110 (35.9) | 19 (17.3) | 91 (82.7) | Reference |  |
| Foreign-born | 196 (64.1) | 90 (45.9) | 106 (54.1) | **0.65 (0.56 – 0.76)** |  |
| Missing | 4 | 2 | 2 |  |  |
| Race/ethnicity |  |  |  |  |  |
|  |  |  |  | Crude PR (95% CI) | Adjusted PR* (95% CI) |
| Black | 151 (49.3) | 74 (49.0) | 77 (51.0) | **1.53 (1.28 – 1.83)** | **1.48 (1.11 – 1.97)** |
| Other race | 155 (50.7) | 34 (21.9) | 121 (78.1) | Reference | Reference |
| *Missing* | 4 | 3 | 1 |  |  |
| Tobacco use |  |  |  |  |  |
| Never smoker | 236 (78.7) | 95 (40.3) | 141 (59.7) | Reference |  |
| Current/Former smoker | 64 (21.3) | 12 (18.8) | 52 (81.2) | **1.36 (1.16 – 1.59)** |  |
| *Missing* | 10 | 4 | 6 |  |  |
| Diabetes mellitus |  |  |  |  |  |
| No | 288 (92.9) | 108 (37.5) | 180 (62.5) | Reference |  |
| Yes | 22 (7.1) | 3 (13.6) | 19 (86.4) | **1.38 (1.14 – 1.67)** |  |
| Body mass index (n=296) |  |  |  |  |  |
| Underweight/Normal | 89 (30.1) | 46 (51.7) | 43 (22.6) | Reference | Reference |
| Overweight/Obese | 207 (69.9) | 60 (29.0) | 147 (71.0) | **1.47 (1.17 – 1.85)** | **1.47 (1.04 – 2.10)** |
| *Missing* | 14 | 5 | 9 |  |  |

| Participants<br>Characteristics | Total<br>population<br>(n=310) | No HTN/Abnormal<br>BP at initial visit<br>n (%) = 111 (35.8) | HTN/Abnormal BP<br>at initial visit<br>n (%) = 199 (64.2) | Measure of Association |  |
| --- | --- | --- | --- | --- | --- |
|  |  |  |  | Crude PR (95% CI) | Adjusted PR* (95% CI) |
| Prior BCG vaccination |  |  |  |  |  |
| No | 129 (41.6) | 33 (25.6) | 96 (74.4) | Reference |  |
| Yes | 98 (31.6) | 41 (41.8) | 57 (58.2) | **0.78 (0.64 – 0.95)** |  |
| Unknown | 83 (26.8) | 37 (44.6) | 46 (55.4) | **0.75 (0.60 – 0.93)** |  |

\*Variables in the full model included age group, sex, country of birth, being Black race, tobacco smoking, diabetes mellitus status, BMI category, prior BCG vaccination, history of intravenous drug use, hyperlipidemia, currently on medication for other diseases, and history of imprisonment; but results were only shown for variables included in the final model selected with the stepwise backward selection
Abbreviations:
BCG – Bacillus Calmette-Guérin; BP – blood pressure; HTN – hypertension; US – United States
**Bold** indicates that the finding is statistically significant with $\alpha=0.05$

Among participants with IGRA results available, 72% (72/100) had the quantitative IGRA measures recorded. Among this subset, we did not observe a significant correlation between quantitative IGRA measures and systolic BP (Supplementary Figure S2, p>0.05). For example, for every unit increase in IGRA nil values, the systolic BP increased on average by 0.632 point (95%CI -4.326 – 5.591) (Supplementary Figure S2A). Furthermore, for every unit increase in TB antigen 1 – nil values, the systolic BP decreased on average by 0.280 point (95%CI -2.289 – 1.730) (Supplementary Figure S2B). Similarly, for every unit increase in TB antigen 2 – nil and mitogen – nil values, the systolic BP decreased on average by 0.971 (95%CI -2.709 – 0.767) and 0.147 (95%CI -0.577 – 0.282), respectively (Supplementary Figure S2C and S2D). We also did not observe a significant correlation between quantitative IGRA measures and diastolic BP (Supplementary Figure S3, p>0.05). For example, for every unit increase in IGRA nil values, the diastolic BP increased on average by 1.34 point (95%CI -2.404 – 5.083) (Supplementary Figure S3A).

Furthermore, for every unit increase in TB antigen 1 – nil values, the diastolic BP decreased on average by 0.389 point (95%CI -1.908 – 1.129) (Supplementary Figure S3B). Similarly, for every unit increase in TB antigen 2 – nil and mitogen – nil values, the diastolic BP decreased on average by 0.569 (95%CI -2.005 – 0.868) and 0.0702 (95%CI -0.389 – 0.249), respectively (Supplementary Figure S3C and S3D).

Our study found that nearly two-thirds of adults with LTBI presenting to a real-world clinic setting in a low-TB endemic area had an abnormal BP reading and/or history of hypertension. These findings are consistent with a recent NHANES report (9), although our estimated prevalence was slightly higher relative to NHANES estimates. This suggests that the magnitude of association between LTBI and hypertension may be different across different sub-group populations, especially those presented in the clinical settings.

Our study findings highlight the need to integrate existing non-communicable disease screening and prevention services with tuberculosis control programs. Notably, many patients presented to the clinic for required TB testing (immigration process, healthcare worker screening, etc.) and did not have reliable primary care. Furthermore, patients with known hypertension that presented to our clinic with elevated BP, may require optimizing their antihypertensive management. Since the duration of preferred LTBI treatment regimens is 3 to 4 months long, TB programs may offer a unique opportunity to aid in the diagnosis and treatment of hypertension and other chronic diseases with subsequent linkage to primary care services for further management.

We also reported common risk factors of any hypertension as reported in NHANES which includes older age, obesity, and Black race. As these key factors were also reported as risk factors of LTBI in the US population (1), it is critical to follow-up these individuals or establish linkages to cardiovascular disease/other chronic non-communicable disease clinics to prevent further progression or more severe form of any cardiovascular events. Future studies need to consider designs and methods to isolate the effect of shared common risk factors for LTBI and hypertension that may partially explain the observed association between LTBI and hypertension in the present study.

Our study subjects to several limitations. First, our study used single visit BP measurements and, therefore, we could not confirm a hypertension diagnosis. This warrants further clinical investigation with larger prospective studies to better characterize BP readings, including comparison between those who were treated for LTBI vs. those who were not. Second, since we relied on medical charts, we did not have access to other predictors of hypertension and LTBI risk that may affect the association between LTBI and hypertension such as stress, family history, diet, and lifestyle. Consequently, our estimates may be slightly distorted from unmeasured confounding effects. Third, we conducted our study in a single public health clinic in Ohio, preventing us to draw any inference to other clinic settings, especially with different population characteristics that may affect the background prevalence of LTBI and hypertension in the population level.

Despite study limitations, our study provides epidemiologic evidence suggesting that hypertension is common among individuals with LTBI. Integrating TB control program with non-communicable care could improve quality and individuals’ access to care to lower the burden of the two diseases through more centralized public health interventions.

## Data Availability

All data produced in the present study are available upon reasonable request to the authors

## DECLARATIONS AND ACKNOWLEDGEMENTS

## Acknowledgments

We thank the personnel and patients at the Hamilton County Public Health, Ohio.

## Data Availability

Data that supports findings of this study can be made available upon request pending review by the local public health clinic.

## Ethical approval and consent to participate

The study was submitted and approved by the University of Cincinnati Institutional Review Board (UC IRB#2021-0060). Individual consent was not required for retrospective access and extraction of existing, de-identified project data.

## Authors contributions

TMS, ADS, MAH: conceptualization, investigation, data analyses, interpretation of results, writing, review, editing, visualization. RE, ADH, LM, KP, MJM, EMU: interpretation of results, review, editing.

