## Supplement Material for "Abnormal Blood Pressure Among Individuals Evaluated for Latent Tuberculosis Infection in a U.S. Public Health Tuberculosis Clinic"

**Epidemiology and Infection**

**Title:**

**Supplementary Material**

**Supplementary Figure S1. Diagram flow to depict the study inclusion of individuals presenting for TBI at a large midwestern US public health clinic, N=310**

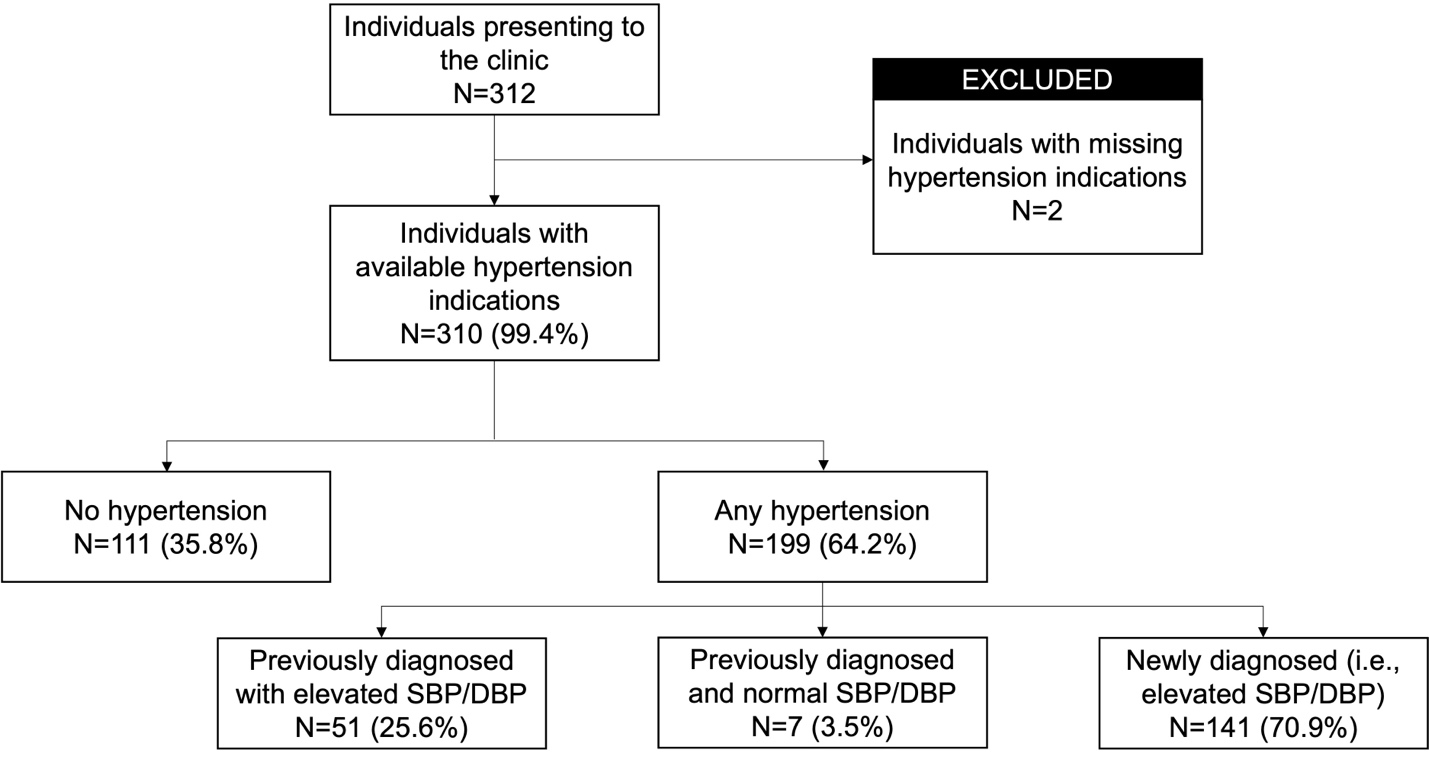

**Supplementary Figure S2. Results from linear regression models to estimate the relationship between systolic blood pressure and A) nil count, B) TB antigen 1 – nil, C) TB antigen 2 – nil, and D) mitogen - nil**

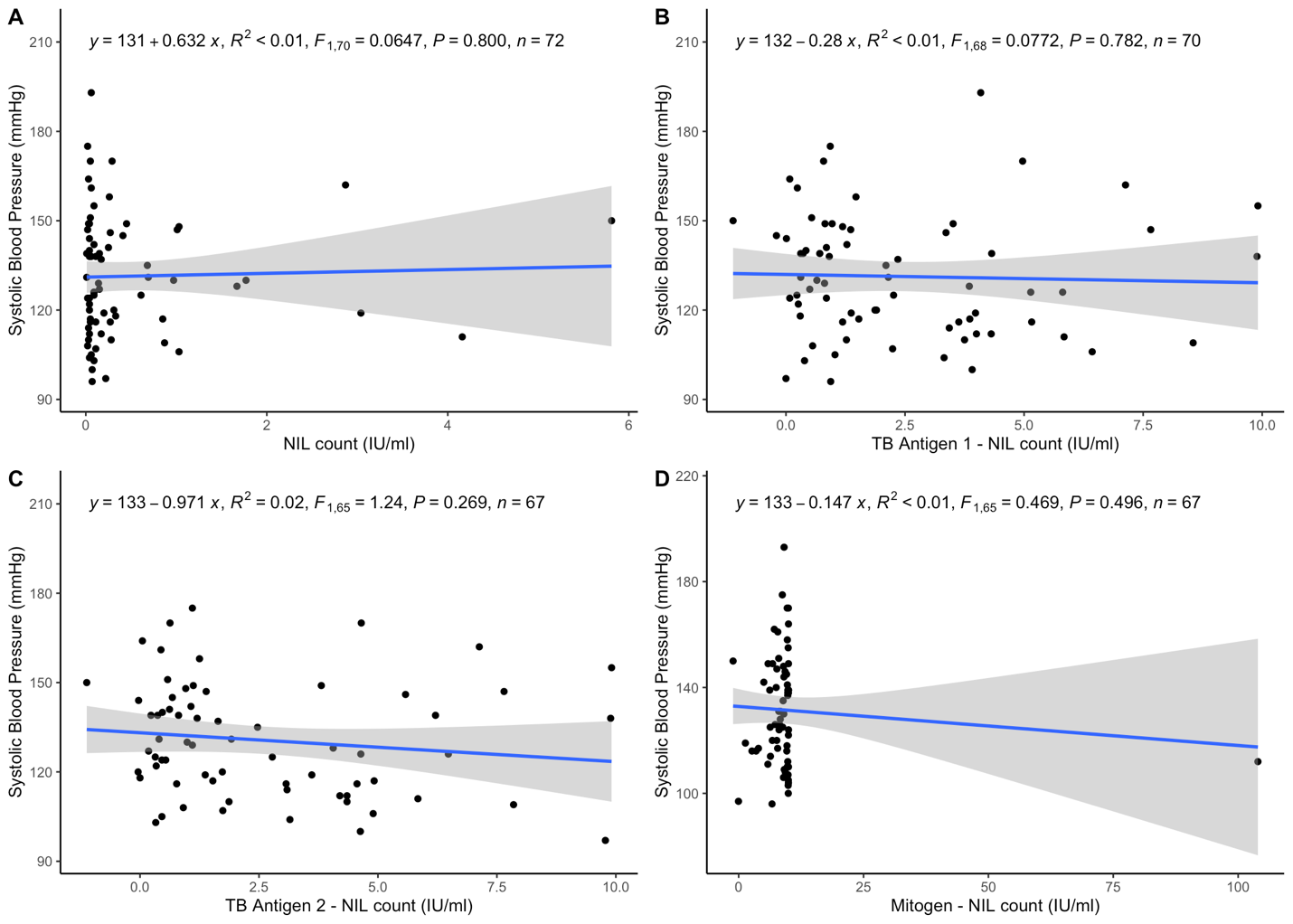

**Supplementary Figure S3. Results from linear regression models to estimate the relationship between diastolic blood pressure and A) nil count, B) TB antigen 1 – nil, C) TB antigen 2 – nil, and D) mitogen – nil**

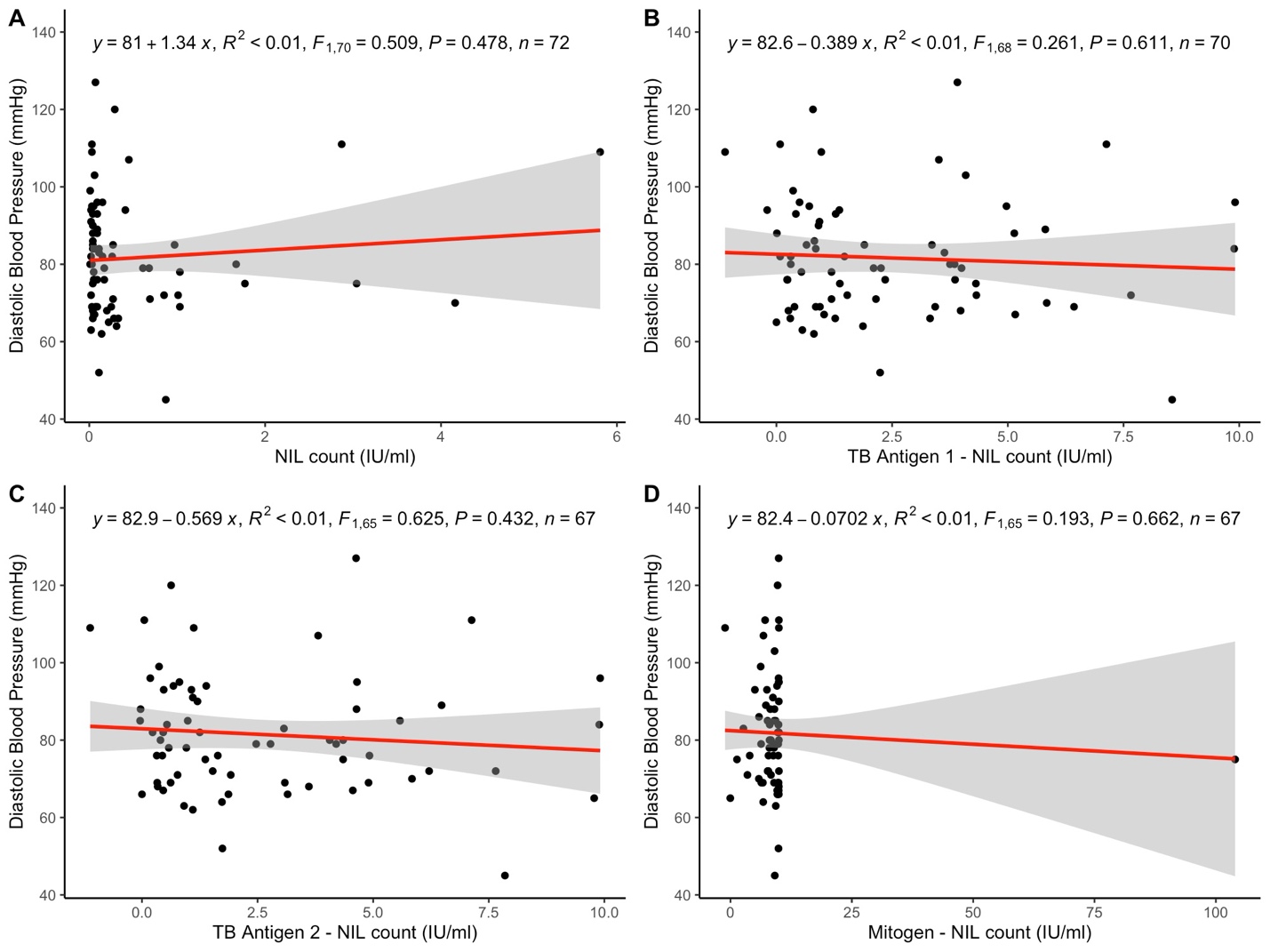
